# Quantifying the heterogeneity and determinants of Ebola Zaïre transmission during the 10th outbreak in DRC, 2018 – 2020

**DOI:** 10.64898/2026.07.14.26358072

**Authors:** Soubrier Hugo, Seo Dohyun, Barks Patrick, Meakin Sophie, Mossoko Mathias, Kitenge Richard, Dierberg Kerry, van Herp Michel, Mambula Christopher, Flasche Stefan, Camacho Anton, Rebecca M. Coulborn, Simons Erica, Ahuka-Mundeke Steve, Broban Anaïs

**Affiliations:** Epicentre, Paris, France; Charité – Universitätsmedizin Berlin, Germany; Médecins Sans Frontières (MSF), Paris, France; Médecins Sans Frontières (MSF), Bruxelles, Belgium; Institut National De la Recherche Biomédicale (INRB), Kinshasa, République Démocratique du Congo; Ministère de la Santé Publique, République Démocratique du Congo; Center for Mathematical Modelling for Infectious Disease, London School of Hygiene and Tropical Medicine, London, United Kingdom

## Abstract

**Background:** The 2018–20 Ebola virus disease outbreak in the Democratic Republic of the Congo (DRC) was the country’s largest, and the second largest globally, amid armed conflict and community mistrust. Transmission heterogeneity (superspreading) is recognised in Ebola epidemics, but empirical estimates of its extent and determinants remain scarce for DRC outbreaks. We quantified transmission heterogeneity and its determinants during this outbreak.

**Methods:** In this retrospective observational study, we reconstructed transmission chains for confirmed and probable cases (Aug 1, 2018, to June 25, 2020) using routinely collected Ministry of Health and Médecins Sans Frontières surveillance data. We modelled the offspring distribution with a Bayesian negative binomial framework, correcting for incomplete contact tracing, to estimate the effective reproduction number (R_eff_), dispersion parameter (*k*), and proportion of cases responsible for 80% of transmission (prop_80_), overall, by subgroup, and over time. Individual-level determinants were assessed with a regression extension, adjusting for covariates.

**Findings:** Among 3481 cases, 2008 transmission events linked 2402 (69%) individuals into 415 chains (median size 3, range 2–102). Overall *R*_*eff*_ was 1.00 (95% CI 0.92–1.08) with *k* 0.29 (0.26–0.32); 17.8% of cases generated 80% of transmission. Overdispersion stayed stable despite fluctuating *R*_*eff*_. Non-isolation (IRR 1.79), death outside a treatment centre (IRR 4.34), and unfollowed contact status (IRR up to 4.48) predicted more secondary cases; vaccination cut transmission by about 60%.

**Interpretation:** Epidemiological investigations linked 67.7% (2356/3481) of cases into 415 transmission chains (median size 3, range 2–102); linkage to a known infector fell to 10% during the November 2018–February 2019 period of peak insecurity. Transmission was heterogeneous overall, with dispersion parameter *k* of 0.29 (0.26–0.32), such that 17.8% (16.8–18.9) of cases generated 80% of onward transmission confirming superspreading as a stable, structural feature of Ebola dynamics. Critically, k remained stable throughout the outbreak, including during periods of elevated *R*_*eff*_, indicating that transmission surges reflected intensification of the same underlying process rather than new superspreading contexts, and that *R*_*eff*_ alone is an insufficient summary of epidemic potential. Regression analyses identified predominantly modifiable determinants: cases not isolated in an Ebola treatment centre (IRR 1.79 [1.54–2.06]) or who died outside one (IRR 4.34 [3.47–5.31]) generated substantially more secondary cases, as did those registered as contacts but not followed up (IRR 3.01 [2.39–3.70]) or unregistered altogether (IRR 4.48 [3.67–5.38]) relative to actively followed-up contacts. Vaccination reduced onward transmission by 60–64% (IRR 0.36–0.40). These findings indicate that transmission was shaped less by gaps in epidemiological knowledge than by the operational reach of contact tracing, isolation, and vaccination delivery, particularly during periods of insecurity.

**Funding:** Médecins Sans Frontières; Einstein-BUA Professorship, Charité – Universitätsmedizin Berlin.

## Introduction

The transmission of Ebola disease (EBOD), a highly lethal haemorrhagic fever, occurs primarily through direct contact with body fluids from symptomatic patients infected with viruses from the *Orthoebolavirus* genus^1,2^. The initial zoonotic spillover and subsequent human-to-human transmission dynamics of Ebola virus arise from complex interactions between environmental, behavioural, and biological factors^3^. Following infection of an index case from an as-yet-unknown reservoir, and in the absence of control measures, sustained human-to-human transmission can occur, resulting in public health emergencies^1^.

To understand transmission dynamics, epidemiological studies have quantified Ebola spread using the basic reproduction number (R_0_), defined as the average number of secondary cases generated by one infected individual in a susceptible population. A comprehensive review of EBOD parameters across reported outbreaks and viral species revealed a highly variable R_0_, ranging from 0.05 to 12^4^ with most estimates derived from data collected during the large West African outbreak (2014-2016). Such variability likely reflects differences in outbreak context, population behaviour, control interventions, and methodological approaches used in estimation. Notably, data from the West African outbreak revealed substantial heterogeneity in transmission, where few individuals infected a disproportionally large number of cases, a phenomenon known as superspreading^5–7^. Evidence from Lau et al. (2017) demonstrated that so-called superspreading events were responsible for sustaining transmission during this epidemic, accounting for up to 60% of secondary infections, and that the extent of superspreading varied in time and place^8^. Subsequent investigations have identified multiple superspreading events, most notably linked to funeral practices^9^, a phenomenon that persisted as recently as the 2022 EBOD outbreak in Uganda^10^. Behavioural factors, specifically the frequency and nature of contact between infected and susceptible individuals largely sustain these transmission mechanisms.

Age^8,11,12^, profession (particularly among healthcare workers)^13,14^, caregiving in the community and hospitals ^6,14,15^, and participation in unsafe burials^9^ are widely recognised as major risk factors for both infection and transmission. Concurrently, biological factors such as disease severity, the presence of wet symptoms (diarrhoea and/or vomiting), and viral shedding likely increase infectiousness15, and persistence of the virus in recovered patients can lead to delayed reactivation years after recovery.^16–18^

In August 2018, the Democratic Republic of the Congo (DRC) experienced its tenth Ebola Virus outbreak, the largest recorded in the country and second largest globally, after the 2014-2016 outbreak in West Africa. The outbreak spanned densely populated urban centres such as Beni and Butembo across the northeastern provinces of Ituri, North Kivu, and South Kivu, and was declared a Public Health Emergency of International Concern (PHEIC) by the WHO on July 17^th^, 2019, following a large increase in cases and evidence of uncontrolled transmission. The response, which involved over forty national and international organisations, was led by the Ministry of Health of DRC, and supported by the WHO^19,20^. This outbreak, caused by the Ebola Zaïre virus (EBOV), was the first to employ a ring-vaccination strategy using the recombinant rVSV-ZEBOV vaccine^21–23^ in combination with the routine core EBOD response pillars known to be effective in reducing transmission and mortality. These included surveillance and contact tracing, clinical management and rapid isolation^24,25^, along with infection prevention and control (IPC) and safe and dignified burial^26^. However, the outbreak was localised in Northeastern DRC, a populous region characterised by a protracted humanitarian crisis where armed-conflict, insecurity, and political instability left the social, economic, and political structures fragile. As a result, the response was undermined by security-related incidents and community mistrust, both of which occurred with increasing frequency and severity, repeatedly leading to delays or temporary suspensions of response activities. Violence culminated in February 2019 following attacks on several Ebola Treatment Centres (ETC) and the temporary suspension of activities by Médecins Sans Frontières (MSF)^20^. The outbreak was declared over on June 25^th^, 2020, nearly two years after its onset, having resulted in 3481 cases and 2301 deaths (case-fatality rate 66.5%).

Estimates of EBOD transmission are highly variable and largely derived from the West African outbreak. Nash et al., (2024) reviewed 32 estimates of the effective reproduction number (*R*_*eff*_), of which only 28% originated from other large epidemics^27^. The extent of superspreading is less well characterised, with 13 estimates of the overdispersion parameter (*k*) reported, of which only two come from DRC epidemics. Additionally, only two studies have leveraged detailed empirical transmission chain data to systematically quantify transmission heterogeneity and its determinants, and none have done so for the 10^th^ DRC outbreak specifically.

Here we address this gap through a retrospective observational study using routinely collected surveillance data from the 10th Ebola outbreak in DRC (2018–2020), from which we epidemiologically reconstructed the largest known set of transmission chains for this disease. We characterized this dataset and the reconstructed chains, quantified key epidemiological distributions, including the offspring distribution and its temporal variation, and identified determinants of transmission.

## Material & Methods

### Data sources

We conducted a retrospective observational study using routinely collected data on confirmed and probable Ebola cases from the tenth outbreak in DRC. These data were collected for clinical purposes as part of the emergency response. The “Master database”, the MoH linelist comprising all confirmed and probable cases, including both individuals admitted for care and those who died in the community, served as our primary data source, from which we matched and merged complementary surveillance databases from the MoH and MSF (details in the Supplementary methods, appendix p. 1).

We integrated transmission chain data independently compiled by MoH and MSF, harmonizing overlapping records and resolving discrepancies as described in the Supplementary methods (appendix page 1). Both datasets were constructed through systematic case investigation and contact tracing, in which field teams linked each case to its most likely infector on the basis of exposure history, setting, and circumstances of contact (e.g. household, healthcare facility, funeral), symptom onset dates, and EBOD’s established 4–21-day incubation period.^28^ Given inherent uncertainties in field-based reconstruction, some cases were linked to multiple plausible infectors while others had no identified source of infection. For cases with multiple plausible infectors, we assigned infectors probabilistically using the serial interval distribution, then used the harmonized transmission chain data to complement the “Master database” by classifying cases according to whether their source of infection was known or unknown, and by quantifying the number of secondary cases generated by each individual.

### Transmission chains

A directed transmission network was constructed from the final transmission-chain dataset (edges = transmission events) and the Master database (nodes = cases in chains), yielding summary datasets on nodes, edges, and chains, while identifying unlinked cases. Chains were defined by size (number of cases), duration (number of days from symptom onset in the index case to symptom onset in the final case), depth (maximum number of transmission generations), and shape (transmission tree topology). Shape was quantified using a branching coefficient, defined as the ratio of chain size to depth, with values in the range 0-1 indicating linear chains and values greater than three indicating star-like topologies.

### Epidemiological delays

We included all cases from the Master database with complete date records for the relevant event pair to estimate the following epidemiological time distributions: isolation delay, reporting delay, admission delay, onset to death interval, onset to recovery interval, length of hospitalisation, and serial interval. All distributions are presented with their observed median and inter quartile range (IQR). More details on the methods used for fitting the distributions can be found in the Supplementary methods (appendix p. 2).

## Model specification

### Offspring distribution model

To estimate key transmission parameters of the EBOD outbreak, we modelled the offspring distribution using a negative binomial framework. We denote the effective reproduction number as *R*_*eff*_, the dispersion parameter as *k*, and define *prop*_*80*_ as the proportion of cases responsible for 80% of onward transmission. The number of secondary cases generated by each case *i*, denoted *V*_*i*_, was assumed to follow a negative binomial distribution with mean *R*_*eff*_ and dispersion 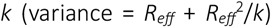, where small values of *k* indicate greater individual-level variation in transmission (overdispersion):

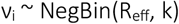

### Correction for incomplete contact tracing

Not all transmission links could be resolved through the contact investigation data. The observed offspring count for each case *i*, denoted n_off,i,_ therefore represents a censored version of the true count *V*_*i*_: only those secondary cases for whom a source case was identified contribute to *n*_*off,i*_. We modelled this observation process explicitly as a binomial thinning of the transmission process, where each true secondary case is independently observed with probability *p*_*B*_ (the probability that a transmission link is identified through contact tracing), which we estimate using the proportion of cases with known infector. Under this two-stage model, the marginal distribution of the observed offspring count simplifies analytically to:

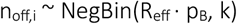

### Transmission chain data

The model was fitted in two stages. First, a global model estimated a single set of parameters (*R*_*eff*_, *k, p*_*B*_) across all cases and the full outbreak duration, providing a summary of average transmission intensity and heterogeneity; *prop*_*80*_ was derived from the joint posterior distribution of *R*_*eff*_ and *k* using the {superspreading} R package^29^. The global model was then refitted independently to subgroups defined by vaccination status and route of contamination to obtain stratum-specific estimates of *R*_*eff*_, *k*, and *prop*_*80*_. For vaccination strata, the *p*_*B*_ prior was informed by stratum-specific linkage counts, whereas for contamination strata the global linkage counts were used as the prior, as all cases within these strata had a known source.

Second, to capture temporal variation, the outbreak period was divided into overlapping rolling 30-day windows (approximately 2.5 mean serial intervals of 12 days) advancing in 1-day steps, estimating window-specific parameters *R*_*eff,τ*_, *k*_*τ*_, and *p*_*B,τ*_ independently. For each window *τ*, the prior on *p*_*B,τ*_ was anchored to linkage counts one serial interval later than the infector window, because whether a secondary case is contact traced depends on response capacity at the time they are detected, not when the infector was symptomatic.

Across all models, we used weakly informative priors on the log-scale transmission parameters: log(*R*_*eff*_) ∼ Normal(log(1), 0.5), centred on the epidemic threshold (*R* = 1), and log(*k*) ∼ Normal(log(0.5), 0.5), centred on moderate overdispersion (*k* = 0.5). The prior on the link-identification probability *p*_*B*_ was specified as a Beta distribution derived from observed linkage counts, Beta(n_linked + 1, n_unlinked + 1); for the global model this corresponded to Beta(1942, 1510), with stratum- and window-specific counts substituted as described above. All parameters were estimated in a Bayesian framework implemented in Stan, with further details provided in the Supplementary methods (appendix p. 2).

### Incidence-based model

In order to compare the estimates of *R*_*eff*_ from the transmission chain model, we inferred *R*_*eff*_ using all the available incidence data and the renewal equation methods provided in the {EpiEstim} R package^30^, applying the same overlapping windows and the empirical serial interval distribution.

### Statistical inference of risk factors

To identify determinants of individual-level transmission, we extended the thinned Bayesian negative binomial model to a regression framework, modelling

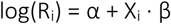

with observed offspring counts again arising from a binomial thinning process. Per-window detection probabilities *p*_*B,τ*_ were fixed as informative Beta priors, moment-matched from the marginal posteriors of the time-varying model described above, so that variation in contact tracing completeness was accounted for without being re-estimated.

Potential exposures were screened for collinearity prior to inclusion, and redundant variables were combined where possible. The following were retained: sex, age group, occupation, health zone type (urban/rural), vaccination status, contact status (not registered; registered but not followed-up; registered and followed-up), chain generation (first case in chain, subsequent case), delay in community (from symptom onset to isolation, or to recovery/death for cases not in isolation), isolation status, final outcome (recovered, death in ETC, death outside ETC), nucleoprotein (NP) CT value, and symptom type (wet: diarrhoea and/or vomiting; dry: absence of gastrointestinal symptoms). Missing data were handled via multiple imputation by chained equations using the R package {mice} (m = 10 datasets; using predictive mean matching)^31^. The regression model was fitted independently on each imputed dataset, and the resulting posterior distributions were pooled to obtain final parameter estimates, propagating imputation uncertainty through the full inference.

A Directed Acyclic Graph (DAG), informed by literature distinguishing biological from behavioural components of transmission^3,7^, was constructed using the R package {dagitty} to derive minimally sufficient adjustment sets for each primary exposure^32^. Only covariates within the relevant adjustment set were included for a given analysis (Supplementary Figure S1). Symptom type and CT value, available only for isolated cases, were examined in a separate sub-cohort analysis. Reference levels were defined as the level with the largest sample size. We report incidence rate ratios (IRR = exp(β)) with 95% credible intervals (95% CI). To avoid “Table 2 fallacies” ^33,34^, that is over-interpretation of adjusted estimates for covariates serving as confounders rather than primary exposures, only estimates for the pre-specified primary exposure are presented alongside their adjustment set. Convergence diagnostics and further sampler settings are described in the Supplementary methods (appendix pg. 4). All analyses were performed in R version 4.3^35^.

## Results

### Datasets

The final Master database consisted of 3481 cases (158 probable and 3323 confirmed). Of these 3481, 2189 (63%) were admitted to an ETC. Most cases (3450, 99.2%) had a valid date of symptom onset. Some data were missing for NP CT value, final outcome, vaccination status, symptom type, occupation, and community delay (details in Supplementary Table S3). The final transmission dataset comprised 2008 transmission events, involving 2402 (69%) individuals from the final Master database. Exposure setting was missing for 12% of these events.

### Epidemiological delays

The delay from symptom onset to case reporting was highly skewed, with a median of six days (IQR 3-9). No consistent pattern in reporting delay was observed when stratifying by health zone or by urban versus rural setting (Supplementary Figure S2 – S3). While the median delay from symptom onset to isolation was four days (IQR 2-7), a decreasing trend in delay to isolation could be seen when stratifying by month of the outbreak (see Supplementary Figure S4). Isolation delay was longest in September 2018 (median: six days), and generally decreased thereafter to a median of three days from October 2019 through the end of the outbreak. Considering all the transmission events recorded, the median serial interval was 12 days (IQR 8-16), and did not meaningfully vary by age group, vaccination status of the infector, chain generation (first vs. subsequent), or exposure setting (Supplementary Table S4). Summaries of other epidemiological delays, and best fit distribution parameters are presented in Supplementary Table S5.

### Epidemiological linkage

Across all cases, 55.8% (1941) had an identified infector, 11.9% (415) had no clear source of infection but generated secondary cases (roots of transmission chains), and 32.3% (1125) remained unlinked. Both in the early and late phases of the outbreak, when fewer cases occurred, more cases had a known infector. The proportion of cases with known ancestor was lowest before the peak of the outbreak between November 2018 and February 2019 (culmination of violence), falling to a low of 10% of cases linked (Figure 1). Linkage rates gradually increased until the end of the outbreak.

**Figure 1.**
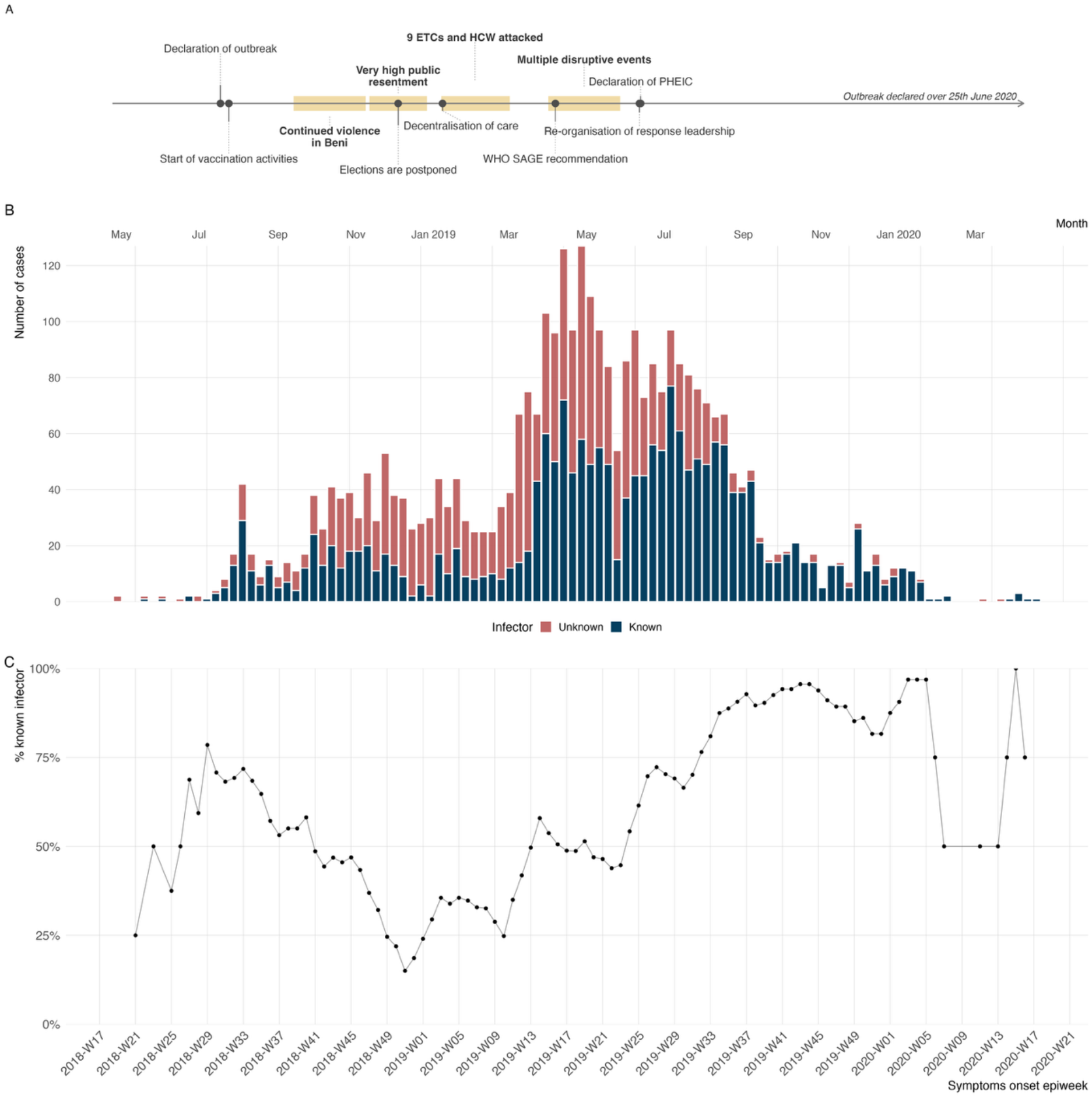
Epidemiological linkage of EBOD cases varied through the outbreak, with a steep decrease in the number of cases linked to an infector between November 2018 and March 2019. A) Simplified timeline of events, highlighted periods of pronounced insecurity and violence are shown in yellow. B) Weekly incidence by symptom onset date stratified by status of infector; unknown (red) and known (blue). C) Rolling average of the proportion of cases linked to an infector over a 4-week period. All panel follow the same x-axis.

### Transmissions chains

Epidemiological investigations linked 2356 (67.7%) individuals into 415 unique chains of transmission. The median chain size was three individuals (range: 2–102), with 179 (43.1%) comprising a single pair of individuals (one infector and one infectee), and 51 (12.3%) comprising ten or more individuals. These large chains accounted for 1176 (33.8%) of all cases. The two largest chains had 90 and 102 nodes. Many chains were short-lived (median duration: 18 days), largely reflecting their small sizes. The 51 largest chains persisted considerably longer (median: 49 days; range: 17–394 days). Geographically, only 13 (25.5%) of these large chains were confined to a single health zone, while 13 (25.5%) spanned four or more health zones and one (2.0%) spanned 12 health zones. Among the 51 largest chains, median depth was four generations (range: 2–8) and 41 (80.4%) had a branching coefficient above 3, indicative of star-like topology characterised by high individual-level secondary case counts. Of the 1941 transmission events recorded across all chains, 1135 (58.5%) occurred in the community, 580 (29.9%) were nosocomial, and 226 (11.6%) were classified as unknown.

### Offspring distribution

Among all cases, 2,513 (72.8%) generated no observed secondary cases. This proportion was lower among the 1,176 cases linked to the largest transmission chains (758; 64.5%). The distribution of secondary cases was heavily right-skewed: 17 individuals (1.4% of cases in the largest chains; 0.49% of all cases) generated more than ten secondary cases, with a maximum of 29.

The estimated effective reproduction number (*R*_*eff*_) was 1.00 [0.92, 1.08], with a dispersion parameter (*k*) of 0.29 [0.26, 0.32], indicating substantial transmission heterogeneity. An estimated 17.8% [16.8, 18.9] of cases accounted for 80% of onward transmission (n = 2,760 secondary cases; Table 1). While an *R*_*eff*_ near unity is expected for a concluded outbreak, the low *k* reveals that transmission was highly concentrated among a small proportion of individuals. Vaccinated cases generated fewer secondary cases but with greater overdispersion than unvaccinated cases or those with unknown vaccination status, such that just 8% of vaccinated cases accounted for 80% of transmission within that group (Table 1). We did not observe differences in transmission between cases vaccinated more or less than 10 days before onset (Supplementary table S6). Cases infected in nosocomial settings had a higher mean number of secondary cases than those infected in community or unknown settings, which both exhibited markedly higher overdispersion (Table 1).

**Table 1.** Estimates of transmission heterogeneity and superspreading (*R*_*eff*_, *k, prop*_*80*_) stratified by infector status.

| Offspring distribution | N | $R_{eff}$ [95% CI] | $k$ [95% CI] | $prop_{80}$ [95% CI] (%) |
| --- | --- | --- | --- | --- |
| **Overall** |  |  |  |  |
|  | 3450 | 1.00 [0.92, 1.08] | 0.29 [0.26, 0.32] | 17.8 [16.8, 18.9] |
| **Vaccination status** |  |  |  |  |
| Unvaccinated | 2308 | 1.26 [1.14, 1.37] | 0.39 [0.34, 0.44] | 21.8 [20.3, 23.3] |
| Vaccinated | 683 | 0.32 [0.24, 0.41] | 0.12 [0.09, 0.17] | 8.0 [6.3, 9.9] |
| Unknown | 459 | 1.39 [1.10, 1.75] | 0.24 [0.18, 0.31] | 16.5 [13.6, 19.7] |
| **Exposure setting** |  |  |  |  |
| Community | 1135 | 0.78 [0.66, 0.91] | 0.20 [0.17, 0.25] | 13.7 [12.0, 15.4] |
| Nosocomial | 580 | 1.41 [1.22, 1.62] | 0.63 [0.48, 0.81] | 27.6 [24.7, 30.6] |
| Unknown | 1735 | 1.01 [0.91, 1.14] | 0.28 [0.23, 0.32] | 17.5 [15.8, 19.0] |

### Temporal variation in transmission

The time-varying effective reproduction number (*R*_*eff,t*_) estimates from transmission data were broadly consistent with estimates from the incidence-based renewal equation approach (Figure 2B). We identified periods of increased transmission in September–October 2018, February–April 2019 and June–July 2019 (Figure 2). On the other hand, overdispersion fluctuated during the first half of the outbreak but subsequently stabilised with 15-20% of cases accounting for 80% of onward transmission (Figure 2).

**Figure 2.**
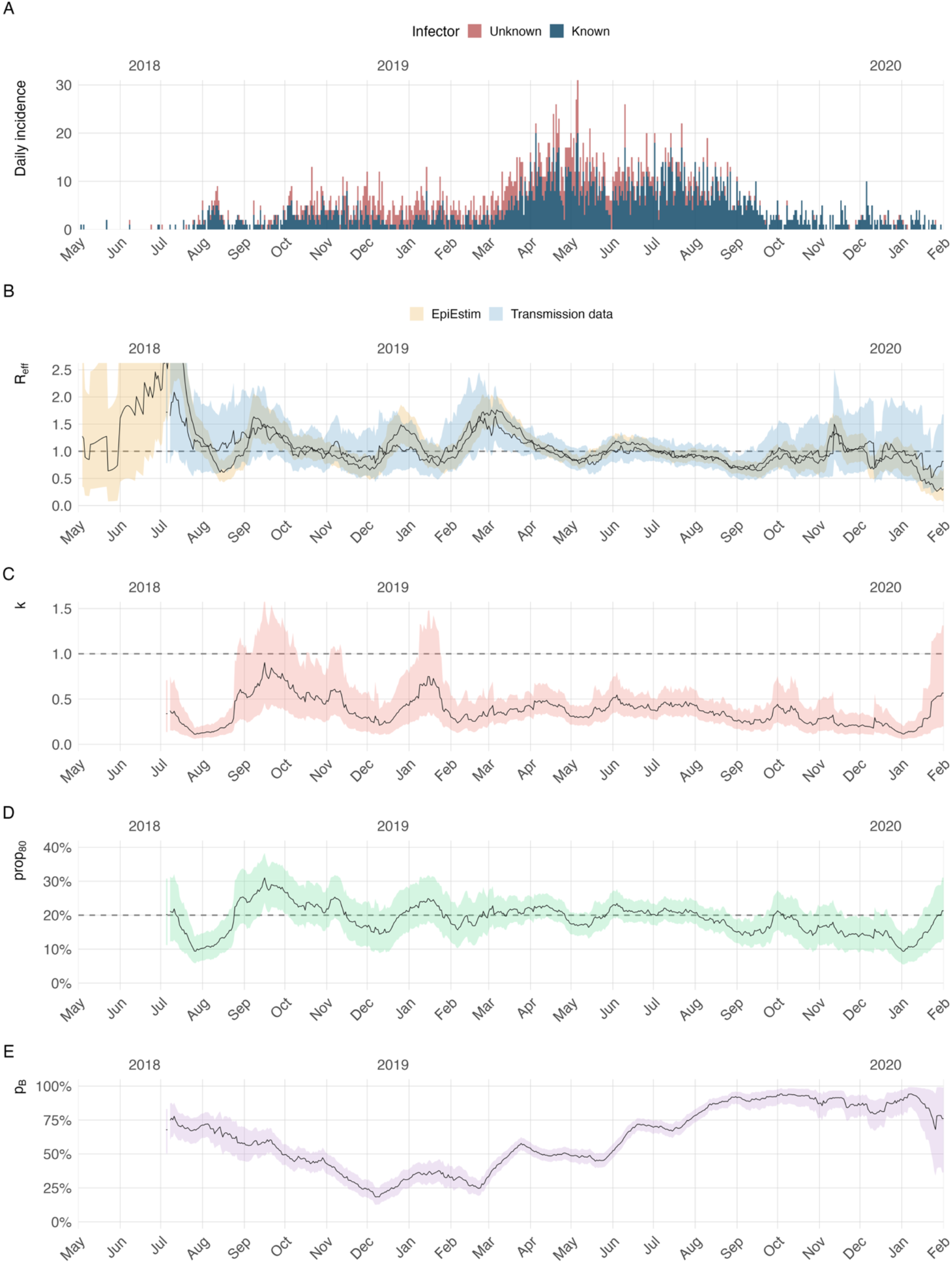
Time-varying transmission dynamics reveal period of sustained transmission and stable heterogeneity across the outbreak. **A)** Daily incidence by symptom onset date stratified by status of infector; unknown (red) and known (blue). **B)** Time-varying effective reproduction number (*R*_*eff*_) estimated from transmission chain data (light blue) and from incidence data (orange). Dotted line indicates *R*_*eff*_ = 1. **C)** Time-varying overdispersion parameter (*k*_*t*_). Dotted line indicates *k* = 1. **D)** Time-varying proportion of cases accounting for 80% of transmission (*prop*_*80*_). Dotted line indicates 20%. **E)** Time varying probability of linkage to source case (*p*_*B*_). In all panels, parameters are estimated over 30 days rolling windows advancing in 1-day steps, from 1 June 2018 to 1 February 2020. Time-windows with sparse data (n < 10) are not plotted. Shaded areas denote the 95% CIs.

### Determinants of transmission

Several variables showed strong evidence of an association with higher numbers of secondary cases after adjustment for minimal sets of confounders (Figure 3). Cases not isolated in an ETC generated 1.79 [1.54, 2.06] times as many secondary cases as those isolated, an effect that was amplified when community delay exceeded four days prior to isolation or end of infectious (Figure 3-delay in community). Among cases that were isolated, low NP CT value was associated with more secondary cases, while having wet symptoms led to 32% more secondary cases than cases with dry symptoms alone (IRR: 1.32 [1.03, 1.65]). Fatal outcomes were associated with higher transmission, an effect markedly stronger among cases who died outside of the ETC (IRR: 4.34 [3.47, 5.31]) compared with cases who survived. Vaccinated cases generated approximately 60% fewer secondary cases than unvaccinated cases, with no observed meaningful difference between cases vaccinated fewer than ten days before symptoms onset (IRR: 0.36 [0.26, 0.48]) from cases vaccinated ten days or more before symptoms onset (IRR: 0.40 [0.18, 0.80]).

**Figure 3.**
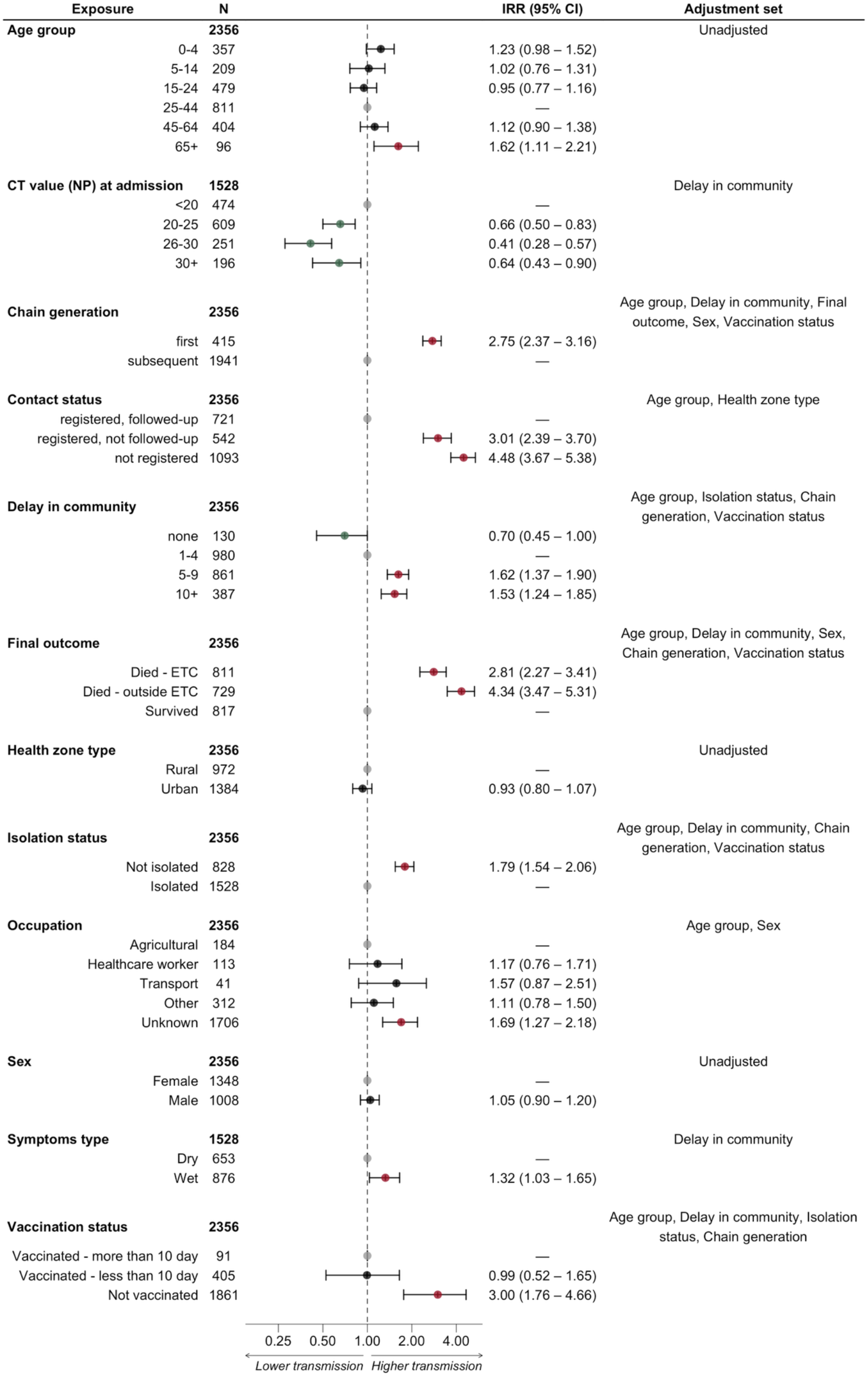
Risk factor analysis of secondary case counts identifies isolation status, final outcome, and EBOD vaccination status as key determinants of transmission. Each row shows the posterior incidence rate ratio (IRR) with 95% credible interval. N denotes the number of cases with non-missing data for each value within each variable). Forest plot shows estimates on the log scale; estimates in red indicate a positive association with secondary case generation (95% CI entirely above 1); estimates in green indicate a negative association (95% CI entirely below 1). Grey points centred on the null (IRR = 1) denote reference levels.

Finally, while being at the root of a chain, and not being registered as a contact were also associated with increased number of secondary cases, we found no evidence of association with occupation, sex, or health zone type (all CIs included 1). No clear pattern emerged between transmission and age, except for cases over 65 years old who generated more secondary cases (1.62 [1.11, 2.21]) than individuals aged 25-44 (Figure 3 – Age group).

## Discussion

This retrospective analysis reconstructed 3,481 cases across 415 transmission chains, producing a detailed characterisation of Ebola transmission dynamics during the 10th DRC outbreak. Three features emerged: transmission was heterogeneous and the level of overdispersion was relatively stable across the outbreak, it was driven by largely modifiable factors. Visibility of EBOD transmission fluctuated with the operational context, highlighting the importance to maintain the response activities that work during the periods when delivering them is most difficult.

Our transmission data revealed marked heterogeneity, confirming that EBOD dynamics are shaped by a small minority of cases that account for a disproportionate share of onward transmission. While more than 70% of cases generated no secondary infections, fewer than 20% accounted for 80% of onward transmission, a pattern consistent with prior estimates from the West African outbreak.^5,7,8^ Rather than being exceptional, superspreading appears to be a fundamental and stable feature of Ebola transmission. Critically, the overdispersion parameter remained relatively stable across the outbreak, including during periods of elevated *R*_*eff*_. This suggests that surges in transmission reflected intensification of the same underlying process rather than the emergence of new superspreading contexts. This finding also challenges the interpretation of *R*_*eff*_ as a sufficient summary of epidemic potential, since transmission can remain sustained even when the mean reproduction number falls below one.^29,36,37^ We therefore echo the recommendation of Nielsen and colleagues (2023) to include the overdispersion parameter (*k*), and its interpretation as the proportion of cases contributing to 80% of transmission, as one of the key epidemiological parameters in EBOD outbreak responses.^36^

Within this overall heterogeneity, transmission in healthcare settings showed a distinct structure. Nosocomial transmission was uniformly elevated, with a higher mean (*R*_*eff*_ = 1.41) and substantially lower overdispersion (*k* = 0.63) than community transmission, consistent with sustained close contact in healthcare settings. Notably, this elevated transmission was not attributable to healthcare workers themselves, who showed no association with higher onward transmission. The relative uniformity of healthcare-associated transmission suggests that standardised infection prevention and control protocols^38^, alongside investment in safe and accessible decentralized care^39^, could reliably reduce transmission in these settings, both directly and indirectly, by preserving the trust required for timely care-seeking when facilities cannot be perceived as safe.

Our analysis provides quantitative evidence that the core pillars of the EBOD response, contact tracing and vaccination, are associated with reduced onward transmission. We observed a clear gradient across contact tracing: cases registered as contacts but not actively followed-up generated 3 (IRR: 3.01 [2.39-3.70]) times more secondary cases than cases that were effectively followed-up, while those unregistered generated 4.48 [3.67-5.38] more. This indicates that the major reduction in onward transmission is driven by active surveillance follow-up. Vaccinated individuals generated 60% fewer secondary cases than unvaccinated ones, an effect that persisted after adjustment for confounding pathways operating through the ring vaccination strategy itself. The persistence of the vaccination effect even after adjustment supports an interpretation of vaccination as a direct protective effect, likely through reduced disease severity and viral shedding, beyond the indirect benefits of the ring strategy.^23^ More broadly, the factors most strongly associated with transmission were behavioural and operational rather than biological, reflecting the frequency and nature of contact between infected and susceptible individuals. These included community delay, absence of isolation in an ETC, death outside an ETC, all reflecting prolonged community exposure of infectious cases or the surveillance gaps that allow it. Biological factors related to virus excretion and disease severity, including CT value and wet/dry symptom profile^2,3^ were also associated with transmission but to a lesser extent. That behavioural determinants dominate is an operationally encouraging finding, since these factors are largely modifiable and are precisely targeted by the core response activities^2,19,23,40^.

In contrast to studies from the West African outbreak, where younger children were less likely to transmit^12^, we did not identify a clear association between transmission and sex or age. The Cellulle d’Analyse en Sciences Sociales (Social Sciences Analytics Cell) (CASS) highlighted that, compared to the West African outbreak, children in the 10^th^ DRC outbreak were less likely to be listed as contacts or followed up systematically, and presented with a different clinical phenotype^38^, both of which could have led to misclassification of transmission events involving young age groups.

Beyond the structure and drivers of transmission, our analysis also characterized how well outbreak responders were able to observe transmission and revealed substantial temporal variation in epidemiological linkage across the outbreak. While more than half of cases had an identified infector overall, fewer than half were registered as contacts at the time of detection, and the probability of linking a case to a known source dropped substantially during periods of operational disruption. These periods of reduced linkage coincided with the surges in transmission described above, driven by attacks on health facilities, population displacement, and community mistrust that compromised contact tracing, case investigation, and vaccination activities.^40–42^ Taken together with the stability of the dispersion parameter described earlier, this pattern indicates that what fluctuated during disrupted periods was the visibility of outbreak responders into transmission. The fragmented structure of many reconstructed chains, often reduced to simple pairs, is consistent with this picture: these fragments likely represent partial reconstructions of a much larger, singly-rooted transmission tree^43^ that could not be fully resolved during the most disrupted periods of the response.

Despite these challenges, investigators and responders succeeded in linking over half of known cases to their infectors and reconstructing 51 large chains comprising hundreds of individuals across multiple health zones over several months. Contact-tracing performance improved over the course of the outbreak itself, with reductions in unlinked cases and isolation delays following the PHEIC declaration, the introduction of decentralized care models^39^, expanded response capacity, and increased international support.^19,20^ Prior analyses have also shown that contact-tracing quality in this outbreak exceeded that of earlier EBOD outbreaks, suggesting that lessons from past responses were effectively applied.^44^

Several limitations should be considered when interpreting these findings. The most important relates to detection bias in chain reconstruction: large transmission bursts are more readily detected through backward contact tracing than smaller transmission events. In our data, first-generation cases in chains show systematically higher offspring counts than later-generation cases, consistent with chains being preferentially identified through their largest transmission events. Those two elements may impact the overdispersion estimate. The binomial correction applied for incomplete contact tracing assumes independent detection probability between cases, an assumption these same observations suggest may not fully hold. Transmission links themselves were inferred from epidemiological data using temporal and contact information, with probabilistic assignment when multiple potential infectors existed, and some misclassification of individual links is likely to have persisted. Missing covariates were addressed through multiple imputation, which reduces but does not eliminate uncertainty in the determinants analysis. Finally, retrospective causal inference from observational outbreak data remains inherently limited: detection biases, unmeasured confounding, and time-varying intervention effects may all influence our estimates and therefore should be interpreted as informative rather than definitive.

## Conclusion

This analysis indicates that transmission during the 10th DRC Ebola outbreak was determined less by gaps in epidemiological knowledge or strategy than by the operational reach of the response. Transmission was heterogeneous and structurally stable, the high-risk profiles were identifiable, and the core response activities, contact tracing and vaccination, measurably reduced onward transmission when delivered. Nevertheless, what fluctuated alongside surges in transmission was the ability of responders to see and act on what was happening. Heterogeneity in transmission emerged as a fundamental and stable feature of EBOD dynamics and should be routinely monitored alongside transmission intensity in future outbreak responses.

The modifiable determinants of transmission share a common mechanism: prolonged community exposure of infectious cases that allow chains to extend unobserved. Operational priorities therefore converge on a small set of activities: community-based alert systems to shorten the time from symptom onset to detection; decentralized case management to increase ETC isolation and reduce community deaths; safe and dignified burial coverage; and sustained contact tracing, particularly the active surveillance follow-up component and vaccination. Protecting these priorities during instability, including by investing in the trust of communities and propose culturally acceptable adaptations where needed, is of the utmost importance to an impactful response.

## Data Availability

All data belong to the Ministry of Health of DRC, who authorised access to the dataset for this collaborative research. Further requests for data access must be approved by the Ministry of Health.

## Funding

This work was funded by Médecins Sans Frontières (MSF) and Einstein – BUA Professorship awarded to Stefan Flasche managed through Charité – Universitätsmedizin Berlin (Grant number: EPP-BUA-2022-697).

## Acknowledgments

The following people were part of the Epicentre-MSF EBOD Working Group and made valuable contributions to data curation and interpretation of the results: Etienne Gignoux, Emmanuel Grellety, Gaston Musemakweli Komanda, Germain Mweha, Rachel Mahamba. The authors are thankful for the work performed by numerous members of the Ministry of Health, Epicentre, MSF, and other partner organisations who contributed to epidemiological investigations that made this analysis possible.

## Data sharing

Code used in the analyses will be made available on Github upon acceptance. All data belong to the Ministry of Health of DRC, who authorised access to the dataset for this collaborative research. Further requests for data access must be approved by the Ministry of Health. Requests should be addressed to Steve Ahuka-Mundeke.

## Ethics

This research fulfilled the exemption criteria set by the Médecins Sans Frontières Ethics Review Board for a posteriori analysis of routinely collected clinical data and was conducted with permission of the Medical Director of MSF Operational Centre Paris. The analysis was also approved by Charité – Universitätsmedizin Berlin review board (reference nº EA1/019/26). The DRC Ministry of Health additionally approved the secondary use of routinely collected programmatic data for our analysis and publication.

## Notes

### Competing Interest Statement

The authors have declared no competing interest.

### Author Declarations

Ethics Review Board of Medecins Sans Frontieres waived ethical approval for this work

