## Supplementary for "Quantifying the heterogeneity and determinants of Ebola Zaïre transmission during the 10th outbreak in DRC, 2018 – 2020"

*Affiliations*

#### Supplementary Methods

##### Data sources

###### *Master database*

To ensure a complete and accurate final master database, we matched and merged surveillance databases of probable and confirmed cases collected during the outbreak as part of the routine activities from the Ministry of Health (MoH) and Médecins Sans Frontières (MSF). The two databases were merged using a unique Identifier, or, when this was missing or unreliable, by probabilistic matching on name, age, and symptom onset date. To retrieve data regarding the isolation, admission and exit date of cases admitted to the ETC, as well as Cycle Threshold (CT) values and type of symptoms at admission, we collapsed the MSF Ebola Treatment Centre (ETC) database to admission records and merged it with the master database, prioritizing ETC-derived values; ETC admission dates were used to define isolation dates. This final dataset was cleaned by recoding categorical variables, resolving implausible or missing dates (including using report date as a proxy isolation date when only ETC records existed), defining outcome dates as death or last discharge for survivors, computing key delay intervals from symptom onset, and setting negative or >50 day delays to missing.

###### *Transmission data*

To ensure consistency and maximize completeness of transmission data, we processed three datasets from MoH (n = 1605, n = 297) and MSF (n = 1551) describing likely infectors assigned for each case by field teams during case investigations. The three datasets were merged and deduplicated using the master database ID. MSF data underwent extensive cleaning, including deduplication, standardization of case identifiers, and extraction of source cases via MSF IDs, VHF codes, or name-based fuzzy matching with the {nmatch} package. For consistency and completeness, cases with multiple potential infectors (n = 147) were resolved by selecting the most probable source using maximum likelihood from a gamma-distributed serial interval distribution estimated from the data; remaining events were validated by removing circular pairs, restricting to master database cases, standardizing contexts of transmission (Nosocomial, Community, Unknown).

#### Epidemiological delays

Delay distributions of epidemiological interest; isolation delay, reporting delay, admission delay, onset to death, onset to recovery, hospitalisation length, and serial interval, were characterised using the {fitdistrplus} package in R[1]. We included all cases with valid temporal information (delays ≥0 and ≤50 days; delays exceeding 50 days were considered data entry errors). Empirical distributions were plotted and summarized descriptively from these valid cases. Candidate parametric distributions (Weibull, Gamma, log-normal) were fitted to the valid data via maximum-likelihood estimation, with the best-fitting model selected by Akaike's Information Criterion (AIC).

#### Model specification

##### *Prior distributions*

**Table S1.** Priors distributions of offspring models

| Model | Parameter | Prior family | Parameters | Justification |
|---|---|---|---|---|
| Offspring distribution models | | | | |
| all Stan models | log(R) | Normal(μ, σ) | μ = log(1), σ = 0.5 | Weakly informative; centred on epidemic threshold (R = 1) |
| all Stan models | log(k) | Normal(μ, σ) | μ = log(0.5), σ = 0.5 | Weakly informative; centred on moderate overdispersion (k = 0.5) |
| Global model | p_B | Beta(α, β) | α = 1942 (n_linked + 1)<br>β = 1510 (n_unlinked + 1) | Derived from observed linkage counts across the full outbreak |
| Stratified — vaccination status | p_B | Beta(α_s, β_s) | α_s = n_linked,<br>s + 1, β_s = n_unlinked,s + 1 (stratum-specific) | Linkage counts within each vaccination stratum |
| Stratified — contamination type | p_B | Beta(α_s, β_s) | α = 1942 (n_linked + 1), β = 1510 (n_unlinked + 1) | Global counts used: all contamination-typed cases have a known transmission source |
| Time-varying model | p_B,τ | Beta(α_τ, β_τ) | α_τ = n_linked,τ + 1<br>β_τ = n_unlinked,τ + 1 (window-specific) | Linkage counts from secondary-case window [τ + 12, τ + 42) days; accounts for detection timing (1 SI) |

##### *Posterior sampling*

All Stan models were fitted via Hamiltonian Monte Carlo using Stan through {cmdstanr}. We ran 4 parallel chains with 1,000 warmup and 2,000 sampling iterations each, yielding 8,000 post-warmup draws. The target acceptance rate was set to 0.95. Convergence was assessed via $\hat{R}$ (threshold < 1.01) and bulk and tail effective sample sizes (threshold > 400). The naive sensitivity model was fitted via rstanarm (2 chains, 2,000 total iterations). Model fit was evaluated using posterior predictive checks comparing replicated and observed offspring distributions.

#### Statistical inference of risk factors

##### *Directed Acyclic Graph (DAG)*

The DAG illustrates the hypothesized causal relationships between demographic, epidemiological, and clinical variables in EBOD transmission. Nodes represent variables, and directed edges indicate causal pathways. Latent (unobserved) variables are shown in grey and include "disease severity" and two latent domains, Biological (related to virus excretion) and Behaviour (frequency and nature of contact between infected and susceptible individuals) as described in Fraser and Grassly (2008).[2]

Observed variables include: age group, sex, vaccination status, urban/rural residence, occupation, contact status, known source of infection (unknown/known), isolation, types of symptoms (wet/dry), community delay to care (delay between symptom onset and isolation or death/recovery), cycle threshold (CT) value at admission and final outcome. The primary outcome, number of secondary contacts infected (n_off), is shown as the outcome node. This DAG was used to identify minimal sufficient adjustment sets for causal inference analyses examining factors associated with EVD transmission while accounting for confounding and selection bias. Arrows indicate presumed direction of causality based on epidemiological knowledge and transmission dynamics of EBOD.

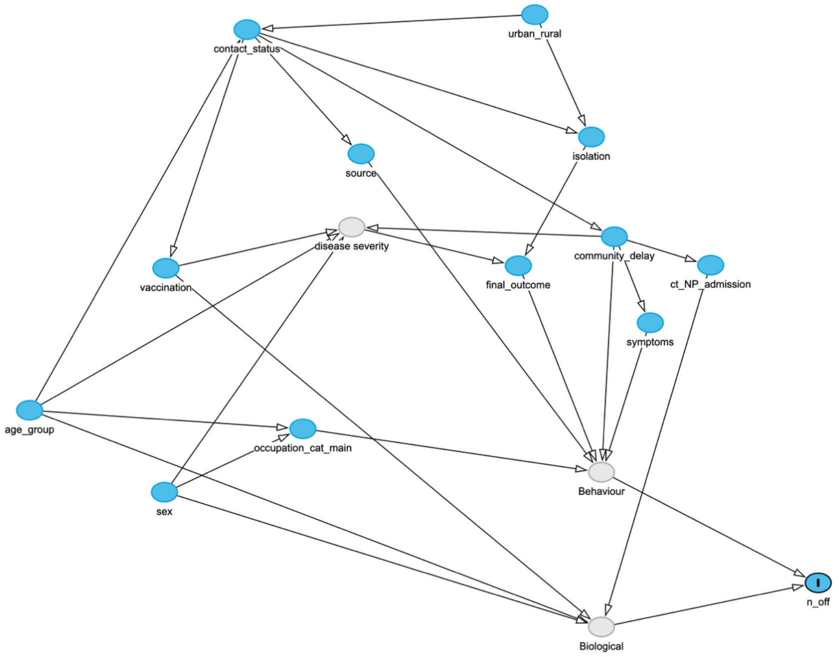


**Figure S1.** Directed Acyclic Graph (DAG) of hypothesised causal pathways for number of secondary EBOD cases (n_off). Latent (unobserved) variables are represented in grey. Available variables in the Master database are represented in blue.

##### *Missing data and multiple imputation*

Missing covariate data were handled using multiple imputation by chained equations (MICE) implemented in the {mice} package. Ten imputed datasets were generated using 30 iterations. The predictor matrix was derived using quickpred() function with the following modifications: onset_date was excluded from all imputation models (it is a passthrough variable used only for window assignment); ct_NP was imputed using a restricted predictor set (n_off, vaccination, community delay, contact status, source, symptoms, urban/rural classification, occupation, sex, and age group) to avoid collinearity with admission-derived variables. The categorical variables ct_NP_admission, symptoms, and community_delay were derived from imputed continuous values after imputation on the long-format stacked dataset, ensuring that categorisation boundaries were applied consistently across all datasets. The final imputed object contained 10 complete datasets each with the full set of predictors and outcome.

##### Variable selection

For each exposure variable, the minimal sufficient adjustment set was derived from the DAG using the {dagitty} package. A separate regression model was fitted for each exposure, including only the exposure and its DAG-derived adjustment covariates. Two cohorts were analysed: the full cohort for all exposures except symptom type and Ct value at admission, which were restricted to the isolated sub-cohort (patients not classified as "not isolated"), as these variables are only meaningful for patients who reached an ETC.

##### *Priors distributions*

**Table S2.** Priors distributions of regression models

| Parameter | Prior | Hyperparameters | Justification |
|---|---|---|---|
| α (intercept) | Normal(μ, σ) | μ = 0, σ = 0.5 | Weakly informative; log(R) ≈ 0 at population-mean covariates |
| β (covariates) | Normal(μ, σ) | μ = 0, σ = 0.5 | Regularising prior; constrains IRR toward 1, reduces collinearity instability |
| k (dispersion) | LogNormal(μ, σ) | μ = log(0.5), σ = 0.5 | Weakly informative; centred on moderate overdispersion |
| p_B,τ (per window) | Beta($\alpha_t$, $\beta_t$) | Moment-matched from time-varying model posteriors | Propagates detection uncertainty from Stage 2 |

##### *Posterior sampling*

Regression models were fitted via Hamiltonian Monte Carlo using Stan through {cmdstanr} on each of M = 10 multiply imputed datasets independently. Each fit used 4 parallel chains with 1,000 warmup and 2,000 sampling iterations, yielding 8,000 post-warmup draws per imputation. The target acceptance rate was set to 0.95. Convergence was assessed per parameter via $\hat{R}$ (threshold < 1.01) and bulk and tail effective sample sizes (threshold > 400). Posterior draws were stacked across all imputations. IRR estimates are reported as the posterior median of exp(β) with 95% highest density intervals (HDI), computed on the pooled draw distribution.

### Supplementary Results

#### Datasets

**Table S3.** Missingness in regression variables prior to imputation

| Variable | N observed | N missing | % missing |
|---|---|---|---|
| Vaccination status | 1,952 | 404 | 17.1 |
| CT value (NP) at admission | 2,193 | 163 | 6.9 |
| Symptom type | 2,255 | 101 | 4.3 |
| Community delay | 2,268 | 88 | 3.7 |
| Final outcome | 2,339 | 17 | 0.7 |
| Age group | 2,356 | 0 | 0.0 |
| Sex | 2,356 | 0 | 0.0 |
| Occupation | 2,356 | 0 | 0.0 |

**Table S3.** Missingness in regression variables prior to imputation

| Variable | N observed | N missing | % missing |
|---|---|---|---|
| Health zone type | 2,356 | 0 | 0.0 |
| Isolation status | 2,356 | 0 | 0.0 |
| Generation in chain | 2,356 | 0 | 0.0 |
| Contact tracing status | 2,356 | 0 | 0.0 |
| Offspring count (outcome) | 2,356 | 0 | 0.0 |

#### Epidemiological delays

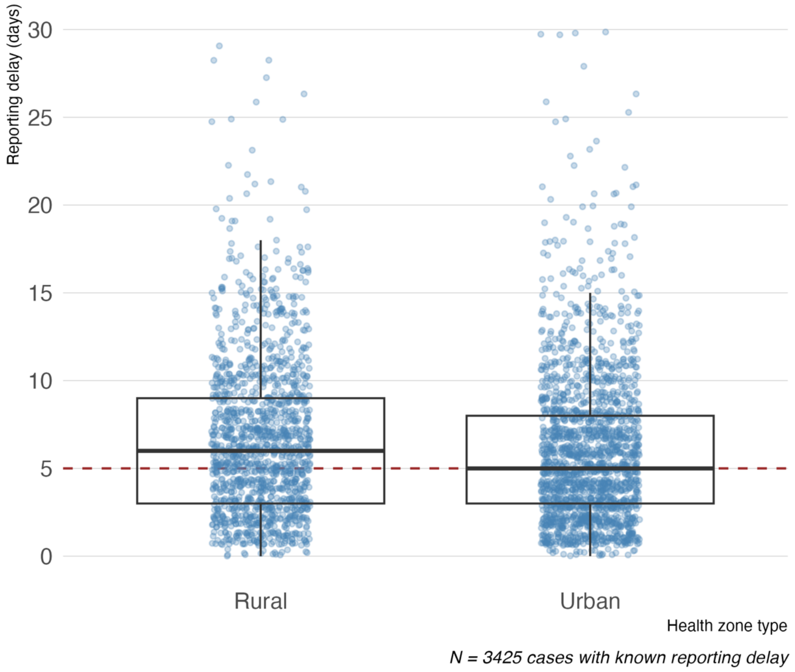


**Figure S2.** Delay between symptom onset and reporting stratified by health zone urbanicity. Red dashed line represents overall median delay (6 days).

**Commented [PB1]:** as below, suggest zooming y axis to (0, 30)

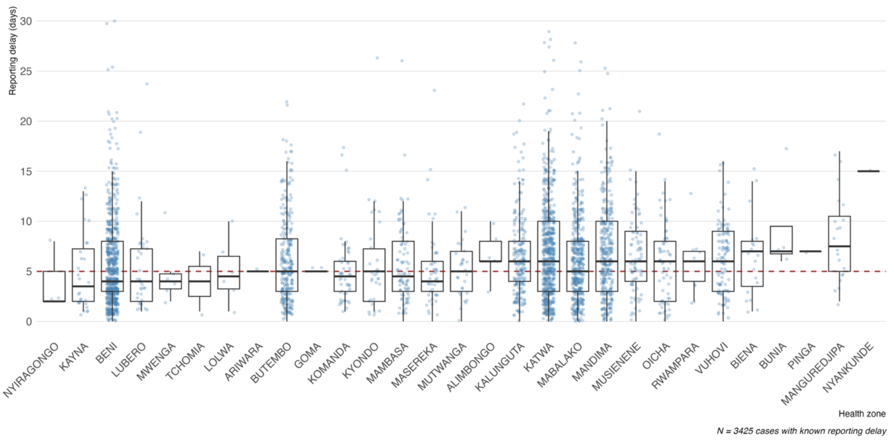


**Figure S3.** Delay between symptoms onset and reporting stratified by Health Zone. Red dashed line represents overall median delay (4 days).

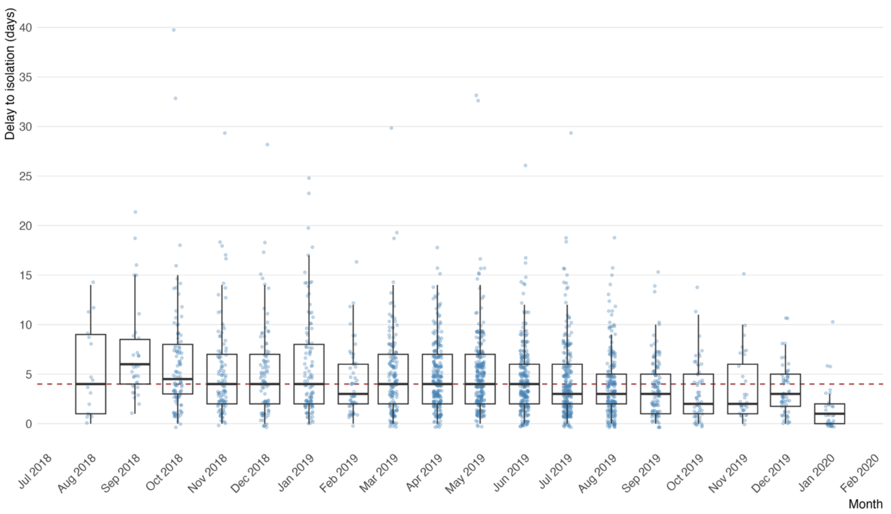


**Figure S4.** Monthly delay between symptoms onset and isolation to an ETC. Red dashed line represents overall median delay (4 days).

**Table S4.** Summary of central tendency and spread for serial intervals, variable refer to the infector.

| | N pairs | Median | IQR | | 95th percentile | |
|---|---|---|---|---|---|---|
| | | | Q25 | Q75 | 2.5th | 97.5th |
| **Age group** | | | | | | |
| 0-4 | 341 | 12.0 | 8.0 | 17.0 | 3.0 | 25.0 |
| 5-14 | 163 | 11.0 | 7.0 | 14.5 | 3.0 | 25.9 |
| 15-24 | 350 | 12.0 | 8.0 | 16.0 | 2.7 | 25.0 |
| 25-44 | 616 | 12.0 | 8.0 | 16.0 | 3.0 | 26.6 |
| 45-64 | 315 | 13.0 | 9.0 | 16.0 | 2.0 | 26.1 |
| 65+ | 111 | 16.0 | 12.0 | 19.5 | 5.8 | 33.8 |
| **Vaccination status** | | | | | | |
| Unknown | 264 | 13.5 | 8.0 | 18.0 | 2.0 | 35.4 |
| Unvaccinated | 1476 | 12.0 | 8.0 | 16.0 | 3.0 | 26.0 |
| Vaccinated | 156 | 10.0 | 7.0 | 14.2 | 3.0 | 20.1 |
| **Exposure setting** | | | | | | |
| Community | 1117 | 13.0 | 9.0 | 17.0 | 3.0 | 27.0 |
| Nosocomial | 571 | 11.0 | 7.0 | 15.0 | 2.0 | 25.0 |
| Unknown | 208 | 12.0 | 7.0 | 16.2 | 1.0 | 29.0 |
| **Chain generation** | | | | | | |
| First generation | 829 | 13.0 | 9.0 | 17.0 | 3.0 | 28.3 |
| Subsequent generation | 1067 | 12.0 | 8.0 | 16.0 | 3.0 | 24.0 |

**Table S5.** Summary of observed delays. Serial interval is presented for the entire dataset, and for the subset of the largest chains (10+ cases).

| | Observed (days) | | | Best-fit distribution | |
|---|---|---|---|---|---|
| Delay | N | Median (IQR) | Range | Distribution | Parameters |
| Onset to reporting | 3343 | 6 (3–9) | 1–92 | Log normal | meanlog = 1.628, sdlog = 0.793 |
| Onset to death | 2159 | 8 (6–11) | 1–44 | Gamma | shape = 3.501, rate = 0.396 |
| Serial interval | 1896 | 12 (8–16) | 1–44 | Weibull | shape = 2.174, scale = 14.236 |
| Serial interval (subset) | 1083 | 12 (8–16) | 1–44 | Weibull | shape = 2.206, scale = 13.818 |
| Admission to death | 1307 | 7 (4–12) | 1–43 | Gamma | shape = 1.739, rate = 0.198 |
| Admission to discharge | 540 | 21 (16–27) | 1–48 | Log normal | meanlog = 2.985, sdlog = 0.487 |
| Onset to admission | 1166 | 3 (2–5) | 1–33 | Gamma | shape = 1.883, rate = 0.451 |
| Onset to recovery | 1113 | 21 (17–26) | 3–65 | Log normal | meanlog = 3.02, sdlog = 0.415 |
| Onset to isolation | 2039 | 4 (2–7) | 1–40 | Log normal | meanlog = 1.318, sdlog = 0.771 |
| Length of hospitalisation | 1857 | 11 (5–18) | 1–66 | Gamma | shape = 1.556, rate = 0.121 |

#### Offspring distribution

**Table S6**. Estimates of transmission heterogeneity and superspreading (*$R_{eff}$, k, $prop_{80}$*) stratified by vaccinated status of the infector.

| Offspring distribution | N | *$R_{eff}$* [95% CI] | *k* [95% CI] | *$prop_{80}$* [95% CI] (%) |
|---|---|---|---|---|
| **Vaccination status** | | | | |
| *Unvaccinated* | 2308 | 1.26 [1.14, 1.37] | 0.39 [0.34, 0.44] | 21.8 [20.3, 23.3] |
| *Vaccinated (less than 10)* | 406 | 0.22 [0.16, 0.31] | 0.19 [0.10, 0.32] | 8.90 [6.7, 11.50] |
| *Vaccinated (more than 10)* | 96 | 0.22 [0.12, 0.36] | 0.55 [0.18, 1.31] | 11.7 [7.5, 16.5] |
| *Vaccinated (unknown delay)* | 181 | 0.76 [0.50, 1.16] | 0.15 [0.10, 0.23] | 11.2 [8.0, 14.7] |
| *Unknown* | 459 | 1.39 [1.10, 1.75] | 0.24 [0.18, 0.31] | 15.6 [13.6, 19.7] |
